# DCM-PROGRESS: predicting end-stage heart failure in non-ischemic dilated cardiomyopathy patients

**DOI:** 10.1101/2023.09.10.23295251

**Authors:** A F Schmidt, P Leinveber, R Panovsky, L Soukup, P Machac, R R van de Leur, A Sammani, K Lekadir, A ter Riele, F W Asselbergs, M J Boonstra

## Abstract

**Aims:** Patients with non-ischemic dilated cardiomyopathy (DCM) are at considerable risk for end-stage heart failure (HF), requiring close monitoring to identify early signs of disease. We aimed to develop a model to predict the 5-years risk of end-stage HF, allowing for tailored patient monitoring and management.

**Methods and results:** Derivation data were available from a Dutch cohort of 293 DCM patients, with external validation available from a Czech Republic cohort of 235 DCM patients. Candidate predictors spanned patient and family histories, ECG and echocardiogram measurements, and biochemistry. End-stage HF was defined as a composite of death, heart transplantation, or implantation of a ventricular assist device. Lasso and sigmoid kernel support vector machine (SVM) algorithms were trained using cross-validation. During follow-up 65 (22%) of Dutch DCM patients developed end-stage HF, with 27 (11%) cases in the Czech cohort. Out of the two considered models, the lasso model (retaining NYHA class, heart rate, systolic blood pressure, height, R-axis, and TAPSE as predictors) reached the highest discriminative performance (testing c-statistic of 0.85, 95%CI 0.58; 0.94), which was confirmed in the external validation cohort (c-statistic of 0.75, 95%CI 0.61; 0.82), compared to a c-statistic of 0.69 for the MAGGIC score. Both the MAGGIC score and the DCM-PROGRESS model slightly over-estimated the true risk, but were otherwise appropriately calibrated.

**Conclusion:** We developed a highly discriminative risk-prediction model for end-stage HF in DCM patients. The model was validated in two countries, suggesting the model can meaningfully improve clinical decision-making.

## Introduction

Patients with non-ischemic dilated cardiomyopathy (DCM) have significant left ventricular (LV) or biventricular dilatation and systolic dysfunction in the absence of abnormal loading conditions, which can result in a major decrease in their quality of life^1^. In up to 30% of DCM patients, a pathogenic mutation can be found, which potentially impacts family members who may be at risk by virtue of their genetic predisposition^2,3^. Risk factors for DCM progression are diverse and include measurements from echocardiography, electrocardiography (ECG), as well as information from patient histories, and patient characteristics such as old age, and low systolic blood pressure (SBP)^4,5^. DCM patients may develop heart failure (HF), and with a 5-year mortality rate of 20%^6^, these patients are at considerable risk for early mortality.

Patients with DCM present with varying signs and symptoms, ranging between early HF symptoms, ventricular arrhythmias or sudden cardiac arrest. This heterogeneous manifestation also translates into substantial differences in progression, where some patients experience improved cardiac function and a relatively sustained quality of life, while other patients are at risk of major adverse events, such as regular hospitalisations or mortality^7^. Treatment for DCM is geared towards management of HF signs and symptoms, as well as prevention or postponement of HF onset. Patients with end-stage HF may be offered a ventricular assist device (VAD) or receive a heart transplant (HTx).

Repeated and close monitoring of these patients forms a cornerstone ensuring DCM patients at risk of disease receive timely preventative interventions. Given the aforementioned differences in disease progression, blanked and stringent monitoring is costly and potentially burdensome. Instead, to allow for more effective and individualized management of DCM we aimed to derive a machine learning model to predict 5-year risk of end-stage HF, a composite endpoint encompassing death, VAD implantation or HTx. We compared performance of our derived *DCM-PROGRESS* model to the MAGGIC score^8,9^ (a European Society Cardiology (ESC) recommended model^10^) predicting mortality in HF patients. Derivation data was sourced from a 293 DCM cohort in Utrecht (the Netherlands), with an external validation cohort consisting of 235 DCM patients from Brno (Czech Republic).

## Methods

### Derivation and external validation cohorts

The derivation cohort consisted of DCM patients referred to the University Medical Center Utrecht (UMCU), the Netherlands, available through the UNRAVEL database^11^. The external validation cohort included patients referred to the International Clinical Research Center (ICRC) of St. Anne’s University Hospital, Brno, Czech Republic. To be considered for the current study, patients needed to have a registered definite clinical diagnosis of non-ischemic DCM, and be 18 years or older. DCM diagnosis was based on: (i) left-or biventricular systolic dysfunction and dilatation or (ii) hypokinetic non-dilated cardiomyopathy with left-or biventricular global dysfunction (left-ventricular ejection fraction (LVEF) less than 45%) which was not sufficiently explained by abnormal loading conditions or coronary artery disease.

The study was conducted in accordance with the Declaration of Helsinki. In UMCU, the study was approved by the local institutional ethics review board (University Medical Center Utrecht, protocol UNRAVEL #12-387). ICRC participants provided written consent for using their data for research purposes, and the study was approved by the local institutional ethics committee (EK-FNUSA-01/2023).

### Candidate predictors

Patient information was extracted from routine care data, based on a 6-months flanking window around the date of enrolment. Enrolment was defined as the referral date if DCM was diagnosed previously, or as the date of DCM diagnosis when determined at or after referral.

The following candidate predictors were available: age (years), female sex, weight (kg), length (cm), SBP, NYHA-class, patient history (life-threatening ventricular arrhythmias [LTVA], non-sustained ventricular tachycardia [NSVT], hypertension, any type of diabetes, atrial fibrillation [AF]; please see the Supplementary for the clinical definitions), family history (cardiomyopathy [CMP], sudden cardiac death [SCD]; please see the Supplementary for the clinical definitions), ECG derived measurements (rhythm AF, QRS duration [ms], heart rate [bpm], QTc time [ms], R-axis (degrees), left bundle branch block [LBBB] morphology, right bundle branch block [RBBB] morphology, paced rhythm [ventricular or atrial]), echocardiographic measurements (LV ejection fraction [EF] in percentage, LV end diastolic internal diameter [EDD] in mm, left atrial (LA) dimension in mm, tricuspid annular plane systolic excursion [TAPSE] in mm, and biochemistry (BNP in pmol/L, NT-pro-BNP in ng/L, creatinine in µmol/L). BNP was mapped to NT-pro-BNP using an established conversion equation^12^.

### Outcome definitions

The study outcome was the 5-years incidence of end-stage HF, defined as a composite of death, implantation of a VAD, or HTx. For the current study, we focussed on events occurring within the first five years from baseline, representing a clinically relevant time-window. To exclude patients referred for immediate intervention (i.e., where there is limited uncertainty in the appropriate treatment strategy), we excluded patients if end-stage HF occurred during the first 30 days of follow-up.

### Feature pre-processing

Missing data were imputed based on a K-nearest neighbour (KNN) algorithm applying a Euclidean similarity matrix^13^ derived using the training data. These training data were subsequently used to prune features which where collinear (based on a pairwise correlation of 0.90), unlikely associated with the outcome (correlation p-value > 0.50), and were invariant (threshold of 0.05). Continuous features were mean-centred and standardized to a standard deviation (SD) of 1; see Supplementary Table S1 describing the training data mean and SD.

### Derivation and validation of DCM-PROGRESS

The UMCU derivation data were randomly split into 80/20% training and testing sets including 234 and 59 DCM patients, respectively. Model hyperparameters (e.g., degree of penalisation) were optimized through 5-fold cross-validation of the c-statistic (i.e., estimating discrimination) stratified by case-control status to ensure a constant outcome incidence in each fold. The training data and cross-validation were used to derive a lasso model (specifically, we optimized a generalized linear model with a binomial distribution and canonical logit link function), as well as a more flexible support vector machine (SVM) model using a sigmoidal kernel. SVM algorithms can naturally account for potential non-linearity and interaction between candidate features^14^. We therefore used SVM as a comparison strategy to determine whether such flexible modelling meaningfully improves predictive performance.

While cross-validation can effectively identify the most appropriate (i.e. most discriminative) hyperparameter, cross-validation is less appropriate to directly compare model performance, especially when applying algorithms with distinct bias-variance trade-offs^15^. As such, the most accurate model was identified through 20% testing sets randomly selected from the UMCU data not used during model derivation. External performance, reflecting potential model transportability, was assessed in the ICRC data.

Performance was evaluated based on discrimination (c-statistic) and calibration (calibration-in-the-large [CIL], and calibration slope [CS]), presented with 95% confidence intervals (CI). We note that under the null-hypothesis of no discrimination, the c-statistic would be 0.50, and a well-calibrated model would have a calibration-in-the-large of 0.00 and a calibration slope of 1.00^16^. Following training, the best performing model was selected based on the largest c-statistic estimated from the hold-out test data, followed by external validation in the ICRC. Feature importance was determined based on the average permuted feature importance, where each feature (i.e., column) was permuted 50 times, breaking any potential association with the outcome. Here the loss in model performance due to permutation, measured in absolute change in c-statistic, reflects the feature’s importance. To unbiasedly determine importance, unaffected by potential overfitting to the training data, the permutation analysis was conducted in the external validation data not used to derive DCM-PROGRESS.

Finally, discriminative performance of DCM-PROGRESS was explored for clinically relevant subgroups of patients with hypertension, patients with a QRS duration longer than 120ms, or patients with a TAPSE larger than 17mm.

### Model recalibration

Lasso and SVM models perform a degree of regularization, which maximizes the c-statistics at a potential cost of decreased calibration (agreement between observed and predicted risk). To correct for this loss of calibration, the training data were used to recalibrate the models after derivation (updating the intercept and slope, see Supplementary note), followed by independent evaluation in the external data.

### Comparisons against the MAGGIC score

Leveraging the external validation data from ICRC, we additionally performed a head-to-head comparison of our novel DCM-PROGRESS model and the MAGGIC score, which has been previously considered in DCM patients^8^. The MAGGIC score was compared to the DCM-PROGRESS based on the aforementioned metrics of discrimination and calibration.

Additionally, we performed a decision curve analysis^17,18^ comparing both models in terms of “net-benefit” against hypothetical scenarios where all DCM patients were offered close monitoring (“Monitor all”) and none is offered close monitoring (“No monitoring”). A decision curve analysis plots net-benefit against a range of probability threshold (here the risk of end-stage HF). Net-benefit is proportional to the proportion of true positives minus the proportion of false positives detected at a certain probability threshold. Of note the theoretical maximum net-benefit is equal to the disease incidence, here 11.5%^18^. The probability threshold can be understood as the number of patients one would be willing to screen to detect one person who will develop end-stage HF. For example, a threshold of 0.10 implies 10 people need to be screened to find one patient who will develop end-stage HF, whereas for a threshold of 0.05 this would be 20. It is important to note that decision curve analyses incorporate both discrimination and calibration, and therefore provides an attractive mode to compare competing models.

### Software to facilitate model deployment and risk communication

We developed an application programming interface (API) facilitating implementation of the DCM-PROGRESS model in care settings. This API additionally includes functionalities to generate graphics relevant for clinical consultation, supporting shared-decision making. Specifically, we developed a function comparing a patient’s estimated risk to a reference risk distribution based on a random 1,000 bootstrap sample of the rounded ICRC data, where a small random error centred around zero was added to ensure anonymity. Additionally, we developed a function to generate a bar chart visualizing the absolute contribution of a patient’s clinical characteristics to the estimated risk. The utility of these graphics will be show-cased using a hypothetical patient with a SBP of 70, r-axis of -5, TAPSE of 25, height of 182, and NYHA class of 1.

## Results

There were 293 DCM patients available in the UMCU data, of whom 65 (22%) developed end-stage HF during a 5-year follow-up period. The ICRC data consisted of 235 DCM patients, with 27 (11%) developing end-stage HF.

The UMCU cohort included more women (110, 38%) and patients had a slightly lower weight (mean 81 kg, SD: 17) compared to the ICRC cohort: 58 (25%) women, mean 92 kg (SD: 20); p-values < 0.01, Table 1. The cohorts additionally differed in terms of patient history (LTVA, hypertension, syncope), family history of CMP and SCD, NYHA class, ECG characteristics (rhythm AF, QRS duration, heart rate, QTc interval, LBBB, pace rhythm), TAPSE, NT-pro-BNP concentration and creatinine concentration; Table 1.

**Table 1.** Patient characteristics at enrolment, stratified by derivation and external validation cohort.

|  | N (%) or<br>Mean (SD) | UMCU (n=293)<br>Median (Q1; Q3) | Missing (%) | N (%) or<br>Mean (SD) | ICRC (n=235)<br>Median (Q1; Q3) | Missing (%) | P-value |
| --- | --- | --- | --- | --- | --- | --- | --- |
| Female sex | 110 (38) |  | 0 (0) | 58 (25) |  | 0 (0) | < 0.01 |
| Weight (kg) | 81 (17) | 79 (70; 90) | 36 (12) | 92 (20) | 90 (78; 105) | 8 (3) | < 0.01 |
| Height (cm) | 176 (10) | 175 (169; 184) | 46 (16) | 176 (9) | 176 (170; 182) | 11 (5) | 0.75 |
| Age (years) | 51 (14) | 52 (42; 61) | 0 (0) | 51 (10) | 51 (44; 58) | 0 (0) | 0.33 |
| History of LTVA | 39 (14) |  | 8 (3) | 11 (5) |  | 0 (0) | < 0.01 |
| History of hypertension | 49 (17) |  | 5 (2) | 125 (53) |  | 0 (0) | < 0.01 |
| History of AF | 38 (13) |  | 0 (0) | 37 (16) |  | 0 (0) | 0.38 |
| Diabetes | 23 (8) |  | 14 (5) | 22 (9) |  | 0 (0) | 0.75 |
| History of NSVT | 21 (7) |  | 0 (0) | 23 (10) |  | 0 (0) | 0.34 |
| History of syncope | 55 (19) |  | 0 (0) | 5 (2) |  | 0 (0) | < 0.01 |
| Family history of CMP | 55 (19) |  | 0 (0) | 21 (9) |  | 1 (0) | < 0.01 |
| Family history of SCD | 24 (8) |  | 0 (0) | 7 (3) |  | 1 (0) | 0.01 |
| NYHA class I | 53 (22) |  | 48 (16) | 23 (10) |  | 9 (4) | < 0.01 |
| NYHA class II | 104 (42) |  |  | 133 (58) |  |  |  |
| NYHA class III | 71 (29) |  |  | 71 (31) |  |  |  |
| NYHA class IV | 17 (7) |  |  | 3 (1) |  |  |  |
| **ECG characteristics** |  |  |  |  |  |  |  |
| Rhythm AF | 46 (16) |  | 4 (1) | 6 (4) |  | 91 (39) | < 0.01 |
| QRS duration (ms) | 113 (28) | 104 (96; 120) | 5 (2) | 127 (36) | 116 (100; 149) | 90 (38) | < 0.01 |
| Heart rate (bpm) | 79 (18) | 77 (67; 90) | 4 (1) | 70 (13) | 69 (61; 78) | 90 (38) | < 0.01 |
| QTc interval (ms) | 455 (46) | 450 (426; 475) | 4 (1) | 433 (38) | 427 (406; 456) | 90 (38) | < 0.01 |
| R-axis (degrees) | 10 (56) | 0 (-33; 41) | 18 (6) | 2 (48) | -6 (-33; 21) | 93 (40) | 0.25 |
| LBBB morphology | 30 (10) |  | 4 (1) | 26 (18) |  | 90 (38) | 0.03 |
| RBBB morphology | 9 (3) |  | 4 (1) | 5 (3) |  | 90 (38) | 1.00 |
| Paced rhythm: atrial | 7 (2) |  | 4 (1) | 6 (4) |  | 90 (38) | < 0.01 |
| Paced rhythm: ventricular | 0 (0) |  |  | 7 (5) |  |  |  |
| **Echocardiographic measurements** |  |  |  |  |  |  |  |
| TAPSE (mm) | 18 (5) | 18 (14; 22) | 123 (42) | 19 (5) | 19 (16; 22) | 47 (20) | 0.02 |
| LA dimension (mm) | 45 (8) | 45 (40; 50) | 174 (59) | 44 (7) | 44 (40; 48) | 5 (2) | 0.40 |
| LV-EF (%) | 27 (10) | 25 (20; 33) | 91 (31) | 27 (9) | 25 (20; 35) | 0 (0) | 0.55 |
| LV-EDD (mm) | 64 (9) | 64 (58; 70) | 81 (28) | 64 (9) | 64 (58; 69) | 9 (4) | 0.58 |
| **Lab measurements** |  |  |  |  |  |  |  |
| NT-pro-BNP (ng/L) | 3230 (6078) | 1002 (418; 2955) | 152 (52) | 2367 (2497) | 1683 (792; 3000) | 94 (40) | 0.02 |
| Creatinine (μmol/L) | 92 (32) | 86 (72; 102) | 116 (40) | 95 (23) | 92 (81; 103) | 79 (34) | 0.02 |

*N.b. The difference between patient characteristics across the two sites were tested using a Mann-Whitney U test for continuous traits, with Fisher's exact test applied to categorical traits. The distributions of continuous traits were summarized using the mean and standard deviation (SD), as well as the median and quartiles (Q) 1 and 3. Categorical traits are described by frequency with percentage. Abbreviations: kg = kilograms, cm = centimeters, bpm = beats per minute, ms = milliseconds, mm = millimeters, LTVA = life-threatening ventricular arrhythmias, AF = atrial fibrillation, NSVT = non-sustained ventricular tachycardia, CMP = cardiomyopathy, SCD = sudden cardiac death, LBBB = left bundle branch block, RBBB = right bundle branch block, TAPSE = tricuspid annular plane systolic excursion, LA = left atrial, LV = left ventricle, EF = ejection fraction, EDD = end diastolic diameter, NT-pro-BNP = N-terminal pro brain natriuretic peptide, heart transplant = HTx, VAD = ventricular assist device, UMCU = University Medical Center Utrecht, and ICRC = International Clinical Research Center.*

### Deriving DCM-PROGRESS to predict 5-years risk of end-stage HF

Pruning candidate features on multicollinearity did not identify variables that needed to be removed (Supplementary Figure S1). RBBB, and paced rhythm were removed due to insufficient variability. Additionally, the features age, history of AF, family history of SCD, QRS duration, diabetes, were removed due to a lack of outcome-association.

The 20 remaining features (Supplementary Table S2) were used to train models predicting 5-year risk of end-stage HF, specifically, we used a lasso as well as a SVM algorithm. The training c-statistic was 0.80 (95%CI 0.74; 0.90) for the lasso model and 0.77 (95%CI 0.68; 0.85) for the SVM model; Supplementary Table S3. The UMCU testing data were used to unbiasedly select the best performing model: where the lasso (c-statistic 0.85, 95%CI 0.54; 0.94) model reached the highest discriminative performance; see Supplementary Table S4. The lasso model was subsequently externally validated in the ICRC cohort, reaching a c-statistic of 0.75 (95%CI 0.61; 0.82); Figure 1. A lasso algorithm performs a cross-validation based feature selection, in this case retaining NYHA class, R-axis, heart rate, height, TAPSE, and SBP; Figure 1, Supplementary Table S5. Permuted feature importance was calculated using the external validation data and described in the Supplementary note and Supplementary Figure 2.

**Figure 1.**
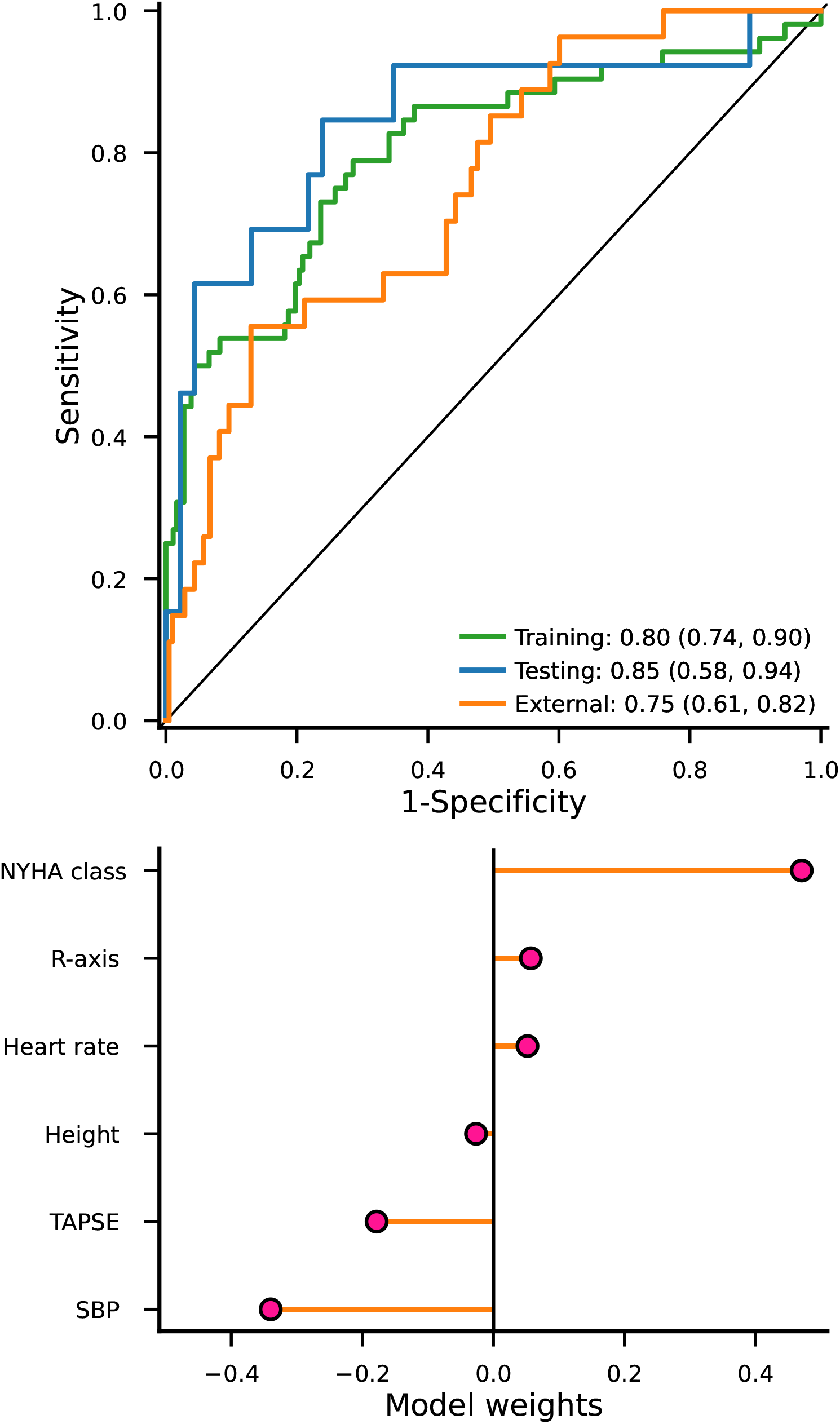
Discriminative performance and model weights of a lasso model predicting end-stage heart failure (a composite of death, HTx, and VAD) in patients with DCM. N.b. heart transplant: HTx, VAD: ventricular assist device, DCM: non-ischemic dilated cardiomyopathy. The c-statistics and 95% confidence intervals are provided as figure annotations, and were estimated using a 1999 iteration bias corrected and accelerated bootstrap. The model weights represent log odds ratio per SD or per class increase for hypertension and NYHA class.

**Figure 2.**
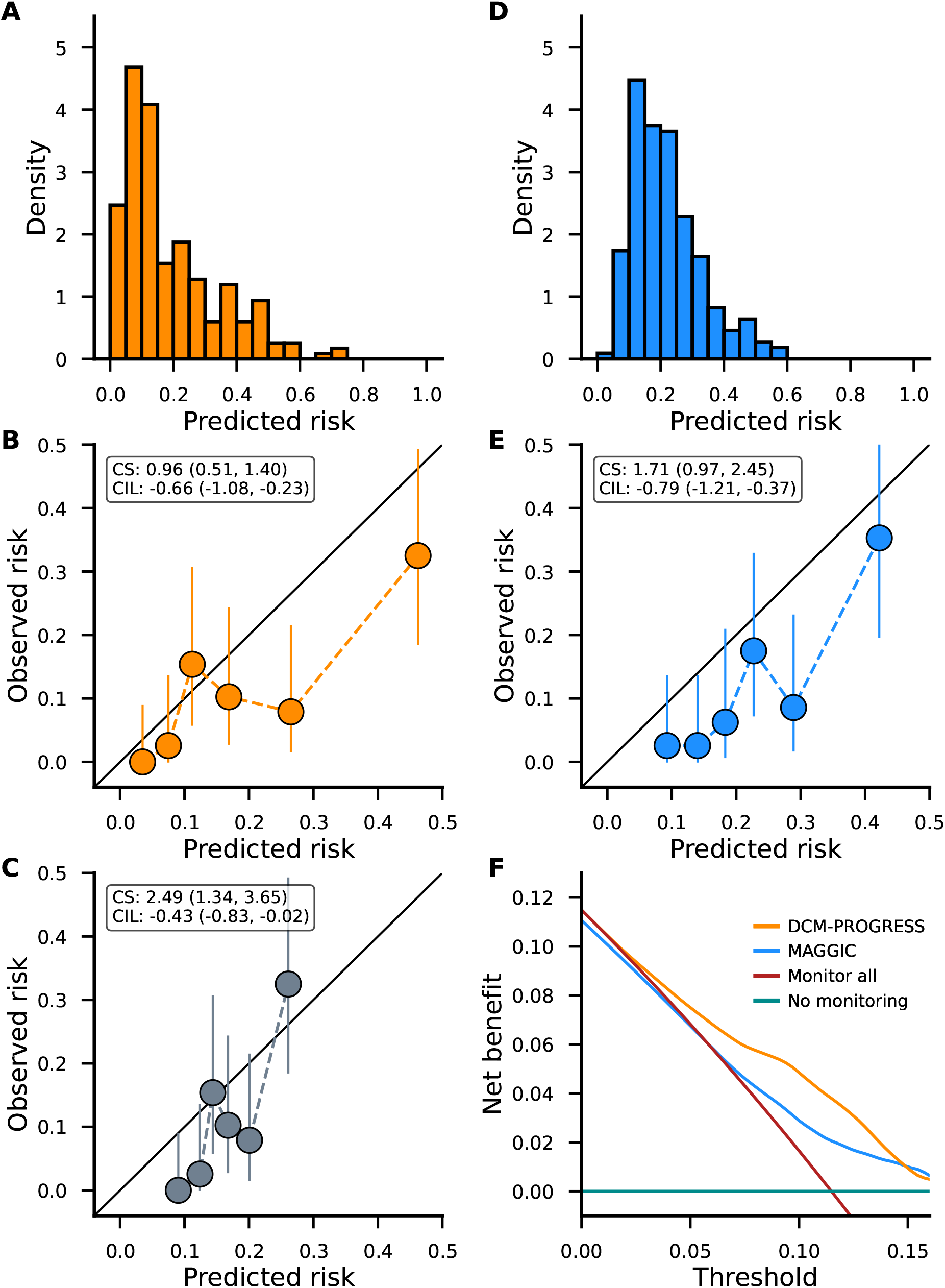
Calibration and decision curve analysis comparing DCM-PROGRESS to the MAGGIC score. N.b. All presented analyses were conducted in the external validation dataset from ICRC consisting of 235 DCM patients of whom 27 developed end-stage HF at 5 years. Panels **A** and **D** present the predicted risk from DCM-PROGRESS and MAGGIC, respectively. Panels **B** and **E**, present depict the model calibration for DCM-PROGRESS and MAGGIC. Note that the DCM-PROGRESS model was first recalibrated (see Supplementary note) using the UMCU training data addressing bias introduced by the lasso algorithm (see methods). For reference performance of the uncalibrated model is included in panel **C**. Based on the predicted risk, participants were grouped into 6 equally size bins, where the x-axis point represents the average bin-specific predicted risk and the y-axis point the mean incidence of end-stage HF. The confidence intervals for the bin specific observed end-stage HF risk were calculated using a beta distribution. Calibration slope and calibration-in-the-large estimates were calculated using the entire dataset without creating bins, with 95% confidence intervals included in brackets. Here a perfectly calibrated model will have a calibration slope of 1.0 and a calibration-in-the-large of 0.0. Finally, panel **F** present a decision curve analysis comparing DCM-PROGRESS and the MAGGIC score in terms of net benefit (i.e., the difference between true positives and false positives). Abbreviations: CS = calibration slope, CIL = calibration-in-the-large.

Subgroup analyses, leveraging the external validation data, indicate that the derived DCM-PROGRESS model performed similarly in participants with hypertension (c-statistic: 0.76 95%CI 0.66; 0.92), QRS duration larger than 120 (c-statistic: 0.73 95%CI 0.53; 0.81), or a TAPSE less than 17 (c-statistic: 0.78 95%CI 0.64; 0.92); Table 2.

**Table 2.** Subgroup performance of the DCM-PROGRESS model predicting 5-years risk of end-stage HF.

| **Subgroup** | **Events/total sample size** | **c-statistic (95%CI)** |
| --- | --- | --- |
| Hypertension | 15/125 | 0.76 (0.66; 0.92) |
| QRS duration (ms) > 120 | 17/68 | 0.78 (0.64; 0.92) |
| TAPSE (mm) > 17 | 18/205 | 0.73 (0.53; 0.81) |

*N.b. The subgroup analyses were performed in the external validation data from ICRC, with 95% bias-corrected and accelerated bootstrap confidence intervals (m = 1999) indicated within brackets.*

### Model calibration

The ICRC data was subsequently used to assess the calibration of the DCM-PROGRESS predicted risk, which ranged between 0.00 and 0.73, and had a median of 0.13; Figure 2A. The DCM-PROGRESS calibration was presented in Figures 2B-C, with Figure 2B presenting the performance of the model recalibrated using the training data (Supplementary note and Supplementary Table S6), and Figure 2C the uncalibrated model. The recalibrated model showed a near-perfect calibration slope (0.96 95%CI 0.51, 1.40), indicating the predicted risk represented the true variability in risk of end-stage HF and accurately predicted risk in the tails of the distribution (i.e., people unlikely versus extremely likely to develop end-stage HF). The estimated calibration-in-the-large (−0.66 95%CI -1.08, -0.23) indicated that the model, on average, slightly over-estimated the predicted risk, implying that the aforementioned median risk is likely slightly overestimated.

### Comparison against the MAGGIC score

The DCM-PROGRESS model was compared to the MAGGIC score using the available external validation data. The estimated c-statistic of 0.69 (95%CI 0.60; 0.82) for the MAGGIC score illustrated that the DCM-PROGRESS model could better discriminate between people with and without end-stage HF in the ICRC cohort. Similar to the DCM-PROGRESS predictions, the MAGGIC score slightly overestimated the predicted risk (calibration-in-the-large -0.79, 95%CI -1.21; -0.37); Figure 2D-E. Unlike the DCM-PROGRESS model, the calibration slope (1.71, 95%CI 0.97; 2.45) indicated the MAGGIC score was unable to accurately predict risk in the tails of the distribution.

Finally, the net-benefit (which jointly assesses discrimination and calibration) of both models was compared across a range of prediction thresholds (Figure 2F). Noting that the maximum net-benefit is equal to the disease incidence (11.5% here), the DCM-PROGRESS model was clearly preferred over a blanked intensified monitoring strategy (even at very low probability thresholds) and outperformed the MAGGIC score.

### Development of a python application programming interface

We have developed an API to facilitate implementation of the DCM-PROGRESS model in clinical care dashboards and potential linkage to electronic healthcare records (https://gitlab.com/SchmidtAF/DCM-PROGRESS). This API is fully tested using modern software development principles such as unit testing, allowing for individual patient calculations (for real-time deployment) as well as bulk calculations (for intermittent deployment). Furthermore, the API contains graphical functionality facilitating shared decision making and risk-communications to individual patients (Figure 3).

**Figure 3.**
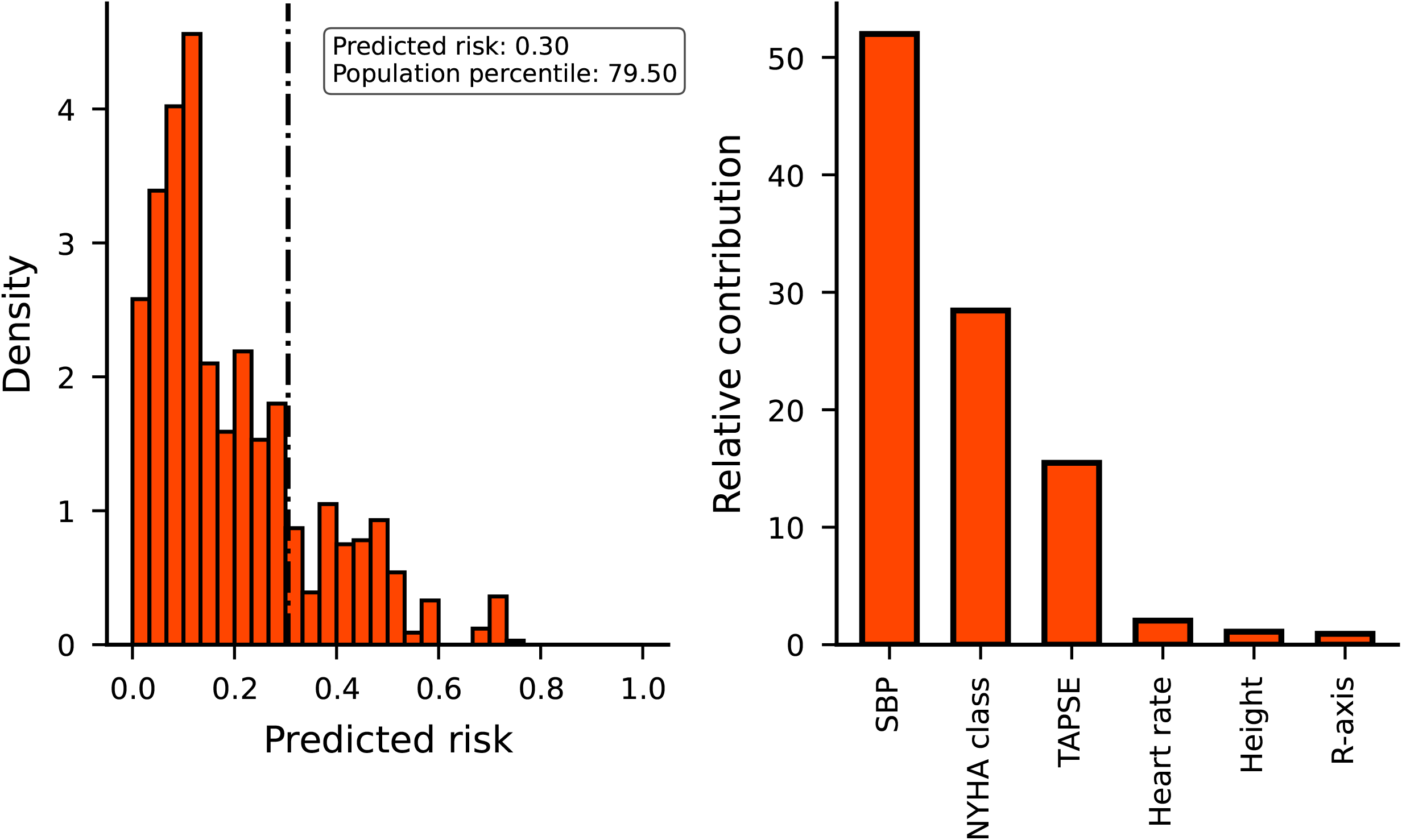
A risk communication illustration comparing a patients estimated risk against a reference population (left panel) and the proportion each clinical characteristics contributed to this risk (right panel) N.b. The left panel plots a hypothetical patients risk as a vertical dashed line on top of a simulated reference population based on the ICRC data (Figure 2A) with their “predicted risk” and “population percentile” annotated. The right panel depicts the relative contribution of the hypothetical patient characteristics to the estimated risk. The risk was estimated based on the following inputs: SBP of 70, r-axis of -5, TAPSE of 25, height of 182 and NYHA class of 1.

## Discussion

In this study we derive a novel clinically applicable prediction model “DCM-PROGRESS”, predicting the 5-years risk of end-stage HF (a composite of all-cause mortality, HTx and VAD) in people with DCM. The model uses six routinely collected variables (NHYA class, R-axis, heart rate, SBP, TAPSE, and heights), and is, therefore readily implementable in clinical practice. Through formal decision curve analysis, we show that DCM-PROGRESS can support clinical decision making in terms of prioritizing patients for intensified monitoring, outperforming a blanketed intensive monitoring of all DCM patients, as well as screening using the MAGGIC sore.

In the current study, we trained and externally validated a prediction model for end-stage HF in people with DCM using two distinct cohorts from UMCU and ICRC. Importantly, we found that despite significant differences in baseline characteristics, as well as differences in the incidence of end-stage HF (Table 1), discriminative performance was similarly robust in both the UMCU hold-out test data and the ICRC external validation data (Figure 1). While both ICRC and UMCU are referral centres for HF and heart-transplantation, the incidence of end-stage HF was higher in UMCU 22% (compared to 11% in ICRC), reflecting differences in patient-groups. For example, at baseline participants enrolled in the two centres differed in terms of proportion of patients with syncope or NYHA class. UMCU typically received patients with more severe DCM, explaining the difference in end-stage HF incidence (Table 1). Through subgroup analyses based on hypertension, QRS-duration, and TAPSE, we confirmed that DCM-PROGRESS reached similar discriminative performances across these clinically relevant patient groups (Table 2). The transportability of the here-derived model, reaching a c-statistic of 0.75 in ICRC data, likely reflects the inclusion of biologically meaningful predictors. For example, TAPSE reflects right ventricular mechanical function, while higher heart rate indicates a compensation mechanism to maintain sufficient cardiac output, with decreased SBP is a well-known indicator of worse prognosis of DCM and HF.

Compared HF risk-prediction models, such as the MAGGIC score, Seattle Heart Failure model^19^ and the Emergency Heart Failure Mortality Risk Grade^20^, DCM-PROGRESS is specifically derived in a non-ischemic DCM-cohort, a patient group often underrepresented. While on average progression in non-ischemic patients is considered more benign compared to progression in ischemic patient, we identified considerable between patient variability, with the predicted risk ranging between 0 and 73%, clearly highlighting the need for (de-)prioritisation of patients for intensified monitoring. As such we expect that implementation of DCM-PROGRESS, can support much needed personalization in patient monitoring and treatment, ensuring the right patient receives the right intervention.

The following potential limitations deserve discussion. Firstly, based on well-established machine learning principles we were able to derive a concise prediction model consisting of six features. We additionally applied a SVM algorithm, which can natively model non-linear or interaction effects, where its lower discriminative performance implies consideration of these type of effects did not improve our model. This lack of increased performance of course does not imply that there are no non-linear or interaction effects, it merely reflects that more complex algorithms typically require more data to derive a model with sufficient out-of-sample performance. The possibility that DCM-PROGRESS might be improved further, does not invalidate the current model, which showed robust predictive performance in both the independent test and external validation samples. Secondly, while the amount of data was limited, it is important to emphasize that in our external validation cohort DCM-PROGRESS outperformed the MAGGIC score in terms of net-benefit, despite the MAGGIC score being derived from a substantially larger number of participants. The performance of DCM-PROGRESS supports the need for DCM specific models, and also illustrates the benefit of applying penalised algorithms such as SVM and lasso, which on average perform better than unpenalized algorithms in relatively sparse data settings. Penalised methods do however decrease calibration^21^ (i.e., agreement between observed and predicted risk), which is why we re-used the training data to re-calibrate DCM-PROGRESS (Figure 2, Supplementary Note). This re-calibration step resulted in a reasonably well-calibrated model, which had a slight tendency to over-estimate the true underlying risk. Through decision curve analysis, calculating the net benefit (i.e., representing the balance between true and false positives) across a range of end-stage HF risk thresholds, we showed that this slight over-estimation of risk did not meaningfully impact clinical utility. This analysis suggested DCM-PROGRESS was a useful tool to decide who is likely to benefit from intensified monitoring, with DCM-PROGRESS prioritisation outperforming both a blanketed intensified monitoring and prioritisation based on the MAGGIC score (Figure 2F). Thirdly, the current study was limited by missing data, which particularly affected echocardiographic measurements and biochemistry measurements such as NT-pro-BNP. Bias due to missing data was minimized through a KNN-based imputation, which due to its non-parametric nature does not induce erroneous linearity of association^13^. Finally, the current 6 feature model is relatively concise and uses information available in most clinical settings. We therefore expect that DCM-PROGRSS can be implemented straightforwardly for example in clinical care dashboards. To facilitate such implementation, we have developed a fully tested and version-controlled python API (https://gitlab.com/SchmidtAF/DCM-PROGRESS), allowing for (real-time and bulk) risk prediction and enhancing patient consultation through graphics presenting the relative risk and risk factors contribution (Figure 3).

In conclusion, we have developed a novel risk-prediction “DCM-PROGRESS” model which predicts the 5-year risk of end-stage HF, and can be used to prioritize patients most likely to benefit from intensified monitoring. Through external validation, we have shown that our model can be transported to new settings, suggesting DCM-PROGRESS can meaningfully support clinical decision-making in DCM patients.

## Data Availability

The derived prediction model is fully described in the supplementary note, with a unit-tested software implementation available on https://gitlab.com/SchmidtAF/DCM-PROGRESS. The underlying patient data is available upon reasonable request and contingent on ethical approval from co-authors Anneline ter Riele (UMCU) and Pavel Leinveber (ICRC).

## Acknowledgments

A preprint version of this manuscript has been deposited at medRxiv.

## Author’s contributions

AFS, MB, FWA, KL, AtR, RP, PL, PM contributed to the idea and design of the study. MB, AS, PL, PM, LS collected and curated the patient data, and AFS performed the analyses. AFS, and MB drafted the manuscript. All authors provided critical input on the analyses and the manuscript.

