## Supplementary Note for "DCM-PROGRESS: predicting end-stage heart failure in non-ischemic dilated cardiomyopathy patients"

**Schmidt *et al.***

**Variable definitions**

| **Variable** | **Definition** |
| --- | --- |
| History of life-threatening ventricular arrhythmia (LTVA) | sudden cardiac arrest, spontaneous sustained VA of >100 bpm lasting >30 seconds or with hemodynamic compromise, ventricular fibrillation or appropriate ICD therapy [either ATP or shock]. |
| History of hypertension | >2 measures of ≥140 mmHg systolic, or use of anti-hypertensive medication. |
| History of atrial fibrillation (AF) | Proven atrial fibrillation during ECG monitoring. |
| History of diabetes | Fasting glucose >7.0 mmol/L OR 2-hour glucose >11.1 mmol/L OR classic symptoms of hyperglycaemia and random plasma glucose of > 11.1 mmol/L. |
| History of non-sustained ventricular tachycardia (NSVT) | three or more consecutive ventricular beats with a rate of >100 beats per minute with the duration of less than 30 seconds without haemodynamic compromise. |
| History of unexplained and/or cardiac syncope | transient loss of consciousness, unexplained or possibly cardiac. |
| Family history for cardiomyopathy (CMP) | at least one 1st and/or 2nd degree family member <65 years of age with proven dilated-, hypertrophic-, or arrhythmogenic cardiomyopathy. |
| Family history for sudden cardiac death (SCD) | at least one 1st degree family member with proven SCD or aborted SCD <50 years of age. |

**Figure S1 A heatmap depicting the pairwise correlation of the 27 candidate features to predict end-stage heart failure in patients with DCM**

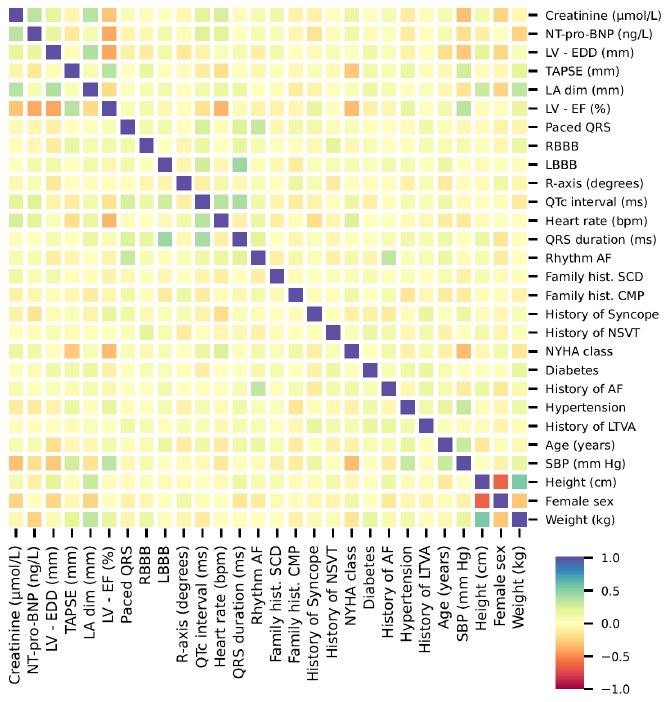

n.b. The Spearman correlation coefficients were calculated using the training data from the University Medical Center Utrecht. The rows and columns have been ordered based on Hamming similarity.

**Implementation of DCM-PROGRESS**

The lasso model included 6 features which jointly predict end-stage heart failure in people with non-ischemic dilated cardiomyopathy: NYHA class, R-axis, heart rate, height, TAPSE and systolic blood pressure. To implement the risk prediction algorithm apply follow the following steps.

*Variable definitions*

NYHA class should be coded as 1, 2, 3, 4. Heart rate should be measured as beats per minute (bpm), R-axis in degrees, height in cm, TAPSE in millimetres (mm), and systolic blood pressure in mmHg. Subsequently the variables should be mean centred and standardized using the estimates in Supplementary Table S1.

*Standard model equation*

Based on the regression weights provided in Supplementary Table S3, the standard uncalibrated risk equation is:

$$\hat{l}=-2.379+0.470\times NYHA+ 0.057\times R-axis+ 0.052\times heart rate-0.027\times height-0.178\times TAPSE-0.380\times SBP (eq 1)$$

Note that for display purposes, the weights are given with 3 decimal point precision, for application please use the full decimal precision provided in Supplementary Table S3.

*Re-calibrated equation*

Given that a lasso algorithm applies a uniform penalization (i.e., it introduces some bias to increase discriminative performance) on all the model coefficients the standard model equations will be sub-optimally calibrated. To correct for this, we re-used the training data to estimate recalibration intercept and slope terms (Supplementary Table S6) which can be used to improve calibration:

$$\hat{l}_{c}=2.572+ 2.604\times\hat{l}.$$

With $l$ defined above and again with the full decimal precision provided in Supplementary Table S6. Importantly given that  $\hat{l}_{c}$ is a rank-preserving transformation of $l$, this step does not affect discriminative performance (i.e., the c-statistic is identical) and merely improves calibration.

*Risk estimation*

The 5-years risk can be estimated using the inverse of the logit function, as follows:

$$\hat{p}=f\left( x \right)=\frac{1}{1+e^{x}},$$

where depending on the desired model $x$ can be substituted by $\hat{l}$ or $\hat{l}_{c}$, and  $\hat{p}$represent the estimated risk (between 0 and 1).

**Feature importance**

The external validation data was used to unbiasedly (i.e., unaffected by potential model overfitting) evaluate the feature importance of the 6 variables included in the lasso model (Figure S2), illustrating that NYHA class, TAPSE and SBP were particularly important features. The negative feature importance (in terms of change in c-statistic) for R-axis and heart rate indicate these variables are likely nuisance features, and do not predict end-stage HF in the external validation data. The presence of nuisance features is generally expected to occur when transporting a model to an external setting, and likely explains the decrease in c-statistic compared to the testing data. It is therefore important to emphasize that despite the presence of some nuisance features, the DCM-PROGRESS model was adequately calibrated, discriminative, and showed increased net-benefit compared to the MAGGIC score.

**Figure S2 Permuted feature importance of the six features used in the DCM-PROGRESS model to predict end-stage heart failure in patients with DCM**

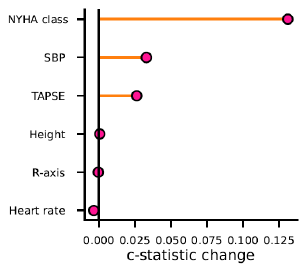

n.b. Feature importance was estimated in the external validation data from ICRC (n=235). Abbreviations: TAPSE = tricuspid annular plane systolic excursion, SBP = systolic blood pressure.
